# Assessment of Cardiovascular Disease Risk Factors in Korean Children: Impact of Various Pediatric Hypertension Guidelines and Application of the Korean Blood Pressure Reference

**DOI:** 10.1101/2024.05.17.24307561

**Authors:** Jeong Yeon Kim, Sangshin Park, Heeyeon Cho

## Abstract

**Background:** The global prevalence of pediatric hypertension (HTN) is increasing and is a significant precursor of cardiovascular disease (CVD). We performed a comparative analysis of two guidelines for pediatric HTN: the 2017 American Academy of Pediatrics (AAP) and the 2016 European Society for Hypertension (ESH); the Korean blood pressure (BP) reference was applied to the Korean pediatric population.

**Methods:** Data from 2,060 children and adolescents aged 10-18 years from the Korean National Health and Nutrition Examination Survey (2016-2018) were analyzed. BP was classified according to the AAP, ESH, and Korea Regional BP Classification (KRC). High BP was defined as BP exceeding the normotensive range.

**Results:** The prevalence of high BP in Korean youth was notably higher in the AAP group than that in the ESH group (19.5% vs. 10.6%, p<0.0001). There were variations in the prevalence based on age, sex, and obesity. No significant disparities were observed between the AAP and KRC groups in terms of high BP prevalence. The application of the AAP and KRC demonstrated a more comprehensive reflection of CVD risk factors, including obesity and metabolic profiles, compared to the ESH. The KRC showed a tendency for more non-obese individuals to be categorized as having elevated BP, although this difference was not statistically significant.

**Conclusions:** This study advocates the adoption of the KRC for defining pediatric HTN in Korea. The KRC identified individuals with CVD risk as having a high BP, which raises the potential of the KRC for early intervention in CVD risk control.

## Introduction

Globally, the increasing prevalence of elevated blood pressure (BP) among youth is becoming a major public health concern.^1–3^ High BP during childhood can lead to early structural or functional changes in vital organs, known as target organ damage (TOD), which may have long-term cardiovascular (CV) implications.^4–8^ The clinical assessment of TOD involves markers such as left ventricular hypertrophy (LVH), microalbuminuria, and etc.^7,8^ Uncontrolled BP elevation increases the prevalence of TOD; however, well-controlled BP can improve this.^9^ While evidence links subclinical TOD in adulthood to clinical CV disease (CVD), the direct association between childhood exposure to CVD risk factors and later-life CVD remains unclear.^10–13^ An ongoing study is evaluating childhood BP levels and predicting future CVD morbidity and mortality.^14^ Similarly, we assessed the consequences of childhood exposure to CVD risk factors based on relevant studies, one of which found that addressing high BP in early childhood improved CVD risk and outcomes in childhood and adulthood.^15–19^ These studies show that childhood exposure to CVD risk factors, such as high BP, increases adult CVD risk.^20^ High BP in childhood is associated with adulthood hypertension (HTN), CVD risk factors, and metabolic syndrome (MS).^21,22^ These are modifiable risk factors for CVD and mortality in adults. Therefore, defining BP elevation in youth is critical for predicting CVD in adults.

Traditionally, the diagnosis of elevated BP in the pediatric population is based on statistical definitions using the distribution of normotensive BPs, as measured by the auscultatory method, according to sex, age, and height in healthy children. We investigated the differences between the American Academy of Pediatrics (AAP) and European Society for Hypertension (ESH) guidelines for diagnosing BP elevation in Korean pediatric populations.^23,24^ These guidelines differ in several ways. The AAP introduced a normotensive BP reference table that excluded data from overweight and obese children to prevent bias from obesity in BP measurements. They also applied static cutoff values from the adult guidelines at a younger age than the ESH. In response to the AAP, Kim et al.^25^ developed a normal-weight BP reference table for Korean youth. However, no BP classification criteria have been proposed for Korean pediatric populations.

In this study, we adapted the AAP BP classification criteria by incorporating a normal-weight Korean BP reference table and creating the Korea Regional BP Classification (KRC). We aimed to evaluate the prevalence of BP elevation in Korean children using various classification criteria and investigate variations in CV risk factor distribution based on these criteria.

## Methods

### Study Population

The data for this study were derived from the Korean National Health and Nutritional Examination Survey (KNHANES) conducted by the Korea Disease Control and Prevention Agency (KDCA) from 2016-2018. The KNHANES adopted a multistage clustered probability design to select its target population, and sample weights were assigned to participants to ensure that the obtained sample accurately represents the broader Korean population. The detailed survey design and data resource profiles are described elsewhere.^26^ We included children aged 10-18 years after excluding those without available height or BP values.

Ultimately, the analysis focused on 2,060 children from the KNHANES dataset to investigate the prevalence of elevated BP (Figure S1). The KNHANES protocol was approved by the Institutional Review Board of the KDCA. Written informed consent was obtained from all the participants and/or legal guardian(s). The present study was approved by the Institutional Review Board (IRB) of Samsung Medical Center (IRB number 21-10-074).

### Clinical and Laboratory Measurements

In the KNHANES, skilled medical personnel conducted anthropometric measurements of participants. Body weight, height, and waist circumference were measured. Age- and sex-specific z-scores were computed for weight, height, and body mass index (BMI) using Lambda Mu Sigma tables from the KDCA.^27^ The z-scores were converted into age- and sex-specific percentiles. Overweight and obesity were defined as a BMI within the 85-94th percentile and ≥95th percentile, respectively, based on the KDCA criteria.

The BP was measured using a mercury sphygmomanometer. Three systolic BP and diastolic BP (DBP) readings were recorded, and final values were obtained by averaging the second and third measurements. BP classification was performed according to the AAP^23^ and ESH^24^ guidelines, using normotensive reference tables from each guideline (Table S1). The KRC, which employs the AAP BP classification using the normotensive reference table from normal-weight Korean youth extracted from Kim et al.,^25^ was also used. High BP was defined as BP greater than the normotensive BP. We additionally analyzed BP according to three age groups: <13, 13-16, and ≥16 years.

Anthropometric and laboratory data were analyzed to assess CV risk factors. Abnormal cutoff points for total cholesterol, high-density lipoprotein (HDL), low-density lipoprotein, triglycerides, fasting serum glucose, and hemoglobin A1c were defined based on the guidelines of the National Institutes of Health and the American Diabetes Association.^28^ MS was defined based on the International Diabetes Federation criteria, which incorporates central obesity and two or more risk factors.^29^ Central obesity was determined using Korean waist reference data.^30^

### CV Risk Factor Profile Differences Between Guidelines

To assess the differences in CV risk factors among children with high BP according to different BP classifications, a comparative analysis was conducted. A subset of 1,799 pediatric participants with complete CV risk factor data was used for this assessment. We compared normotensive individuals to those with high BP according to each criterion. Specifically, among individuals classified as normotensive according to one criterion, those newly classified as having high BP when different criteria were applied were compared with those who remained normotensive. Participants newly diagnosed with high BP by each set of criteria were categorized as “upwardly reclassified” or “upwardly high,” while those consistently normotensive were labeled as “consistently normotensive.” The groups classified as upwardly reclassified/high were compared with age-, sex-, and height-matched consistently normotensive individuals (Tables S2-S4).

### Statistical Analyses

Weighted values were used for the demographic and clinical characteristics of the study population, as recommended by the KNHANES. Continuous variables were presented as weighted mean and standard error, and categorical variables were expressed as weighted percentages with a 95% confidence interval.

The prevalence of high BP according to various BP classifications was compared using the weighted McNemar’s test of symmetry. When comparing upwardly reclassified/high individuals with consistently normotensive individuals, a propensity score analysis was used to match age, sex, and height (percentile) to minimize their potential influence on BP values.

Unweighted paired analyses were used for the comparisons. Continuous variables were presented as mean and standard deviation, and differences were assessed using the paired t-test or Wilcoxon signed-rank test based on normal distribution confirmation. Categorical values were expressed as percentages, and comparisons were made among groups using McNemar’s test.

Statistical analyses were performed using SAS version 9.4 (SAS Institute, Cary, NC, USA), with p-values <0.05 considered statistically significant.

## Results

### Characteristics of the Study Participants

This study examined 2,060 participants aged 10-18 years. The participants had a mean age of 14.3 years; 52.1% were male. Overweight and obesity rates were 9.9% and 12.5%, respectively, and central obesity was present in 10%. The prevalence of MS was 4.2%. Additional demographic and clinical characteristics of the patients are shown in Table 1.

**Table 1.**
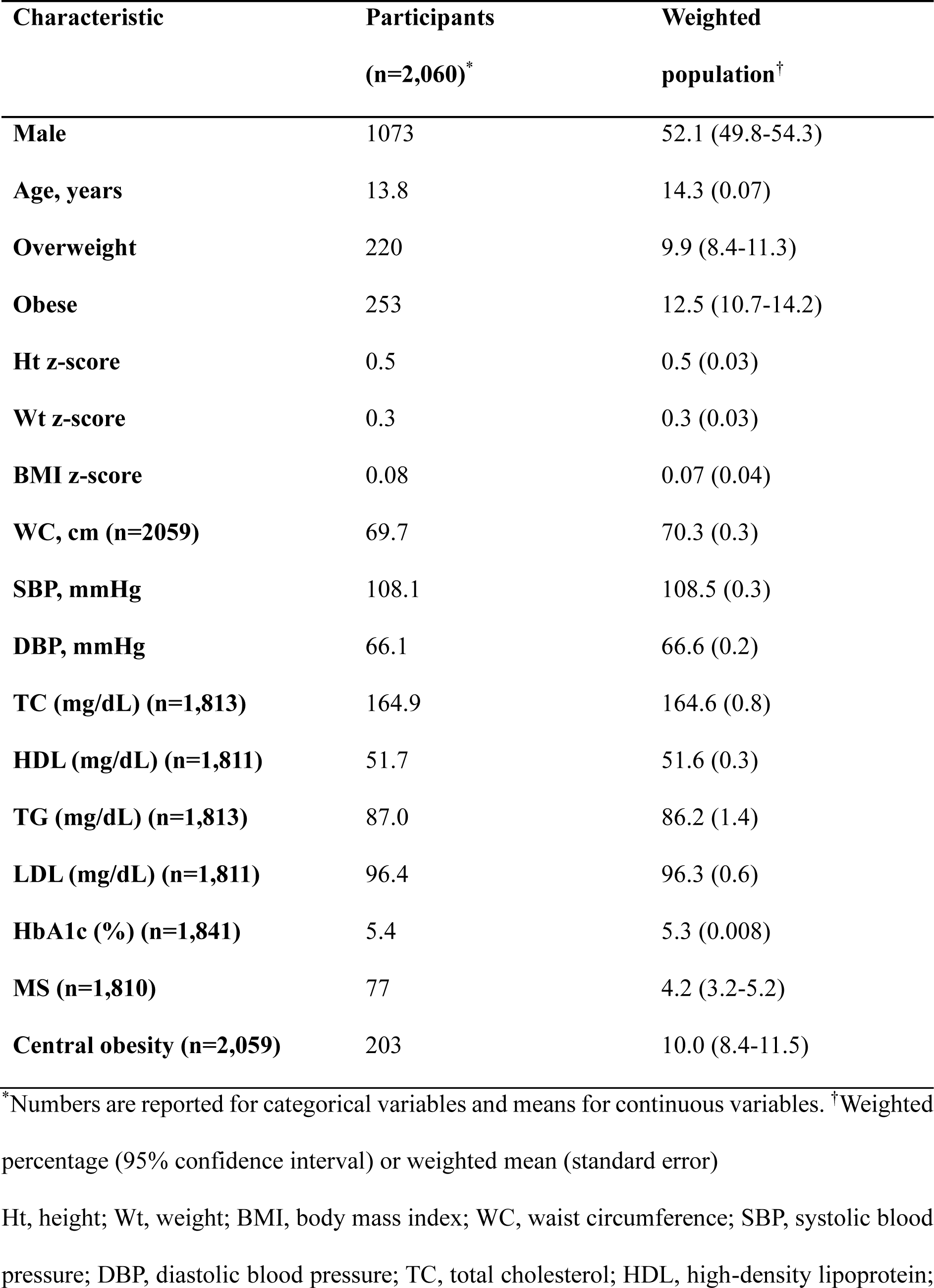
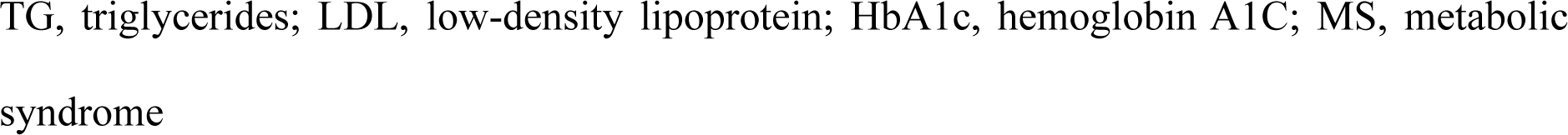
Demographic and clinical characteristic of study population, weighted.

| **Characteristic** | **Participants**<br>**(n=2,060)\*** | **Weighted**<br>**population<sup>†</sup>** |
| --- | --- | --- |
| **Male** | 1073 | 52.1 (49.8-54.3) |
| **Age, years** | 13.8 | 14.3 (0.07) |
| **Overweight** | 220 | 9.9 (8.4-11.3) |
| **Obese** | 253 | 12.5 (10.7-14.2) |
| **Ht z-score** | 0.5 | 0.5 (0.03) |
| **Wt z-score** | 0.3 | 0.3 (0.03) |
| **BMI z-score** | 0.08 | 0.07 (0.04) |
| **WC, cm (n=2059)** | 69.7 | 70.3 (0.3) |
| **SBP, mmHg** | 108.1 | 108.5 (0.3) |
| **DBP, mmHg** | 66.1 | 66.6 (0.2) |
| **TC (mg/dL) (n=1,813)** | 164.9 | 164.6 (0.8) |
| **HDL (mg/dL) (n=1,811)** | 51.7 | 51.6 (0.3) |
| **TG (mg/dL) (n=1,813)** | 87.0 | 86.2 (1.4) |
| **LDL (mg/dL) (n=1,811)** | 96.4 | 96.3 (0.6) |
| **HbA1c (%) (n=1,841)** | 5.4 | 5.3 (0.008) |
| **MS (n=1,810)** | 77 | 4.2 (3.2-5.2) |
| **Central obesity (n=2,059)** | 203 | 10.0 (8.4-11.5) |

\*Numbers are reported for categorical variables and means for continuous variables. <sup>†</sup>Weighted percentage (95% confidence interval) or weighted mean (standard error)
Ht, height; Wt, weight; BMI, body mass index; WC, waist circumference; SBP, systolic blood pressure; DBP, diastolic blood pressure; TC, total cholesterol; HDL, high-density lipoprotein;
TG, triglycerides; LDL, low-density lipoprotein; HbA1c, hemoglobin A1C; MS, metabolic syndrome

### Prevalence of BP Elevation

The prevalence rates of elevated BP/high normal, stage 1 HTN, and stage 2 HTN were 9.4%, 9.3%, and 0.8%, respectively, using the AAP; 6.3%, 3.9%, and 0.4%, respectively, using the ESH; and 9.3%, 9.4%, and 0.8%, respectively, using the KRC (Figure 1A). High BP, as classified by the AAP and KRC, was more prevalent in boys (Figure 1B). Overweight/obese participants exhibited a higher prevalence of high BP for all criteria than did participants with normal weight (Figure 1-C).

**Figure 1.**
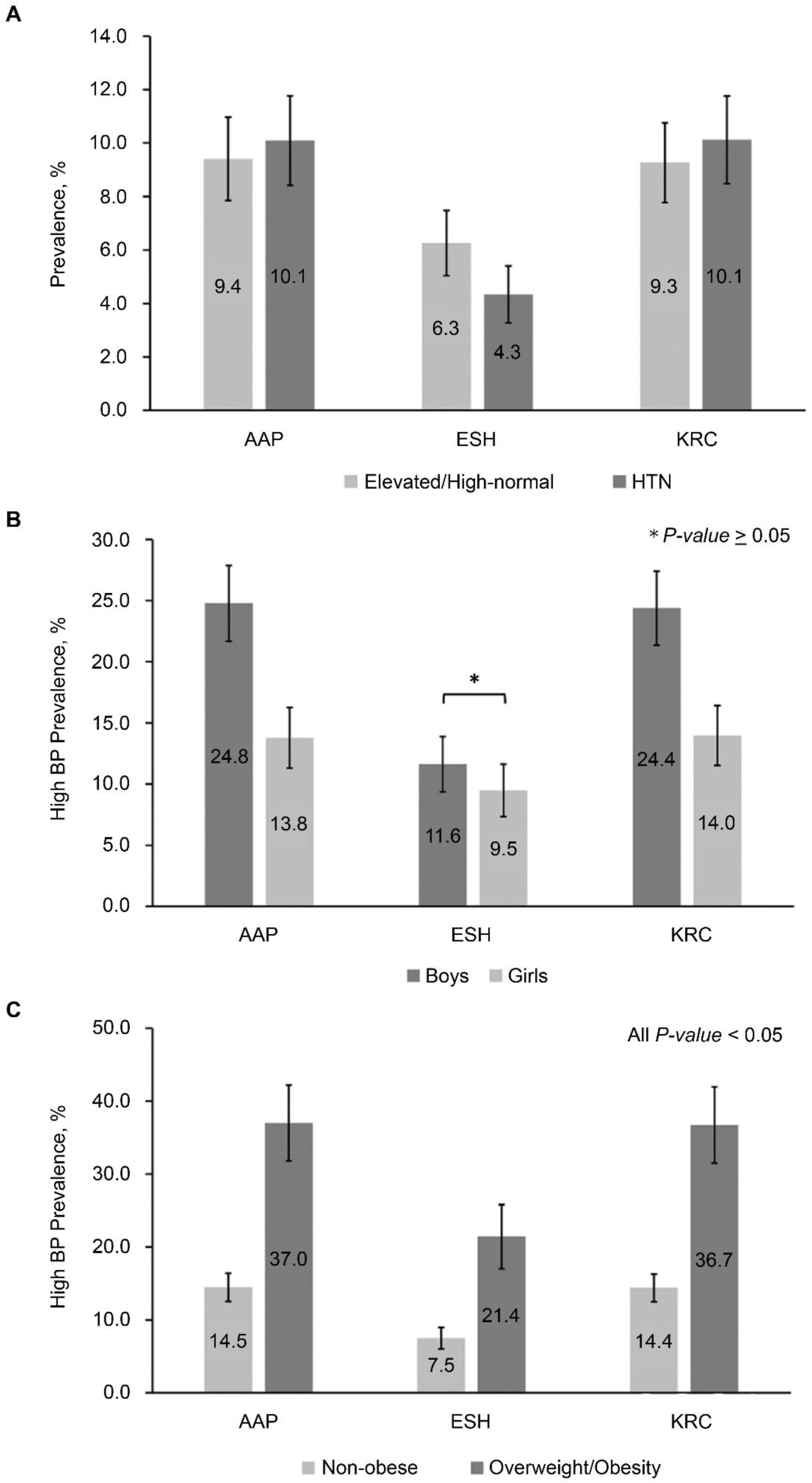
Prevalence of BP elevation in Korean children according to A) each guideline, B) sex, C) obesity, weighted. AAP, American Academy of Pediatrics; BP, blood pressure; ESH, European Society for Hypertension; HTN, hypertension; KRC, Korea Regional Blood Pressure Classification

When data were analyzed according to the three age groups, stage 1 HTN was more prevalent in the <13-year-old and 13-16-year-old groups by both the AAP and KRC than that by the ESH. In the ≥16-year-old group, all BP stages were more prevalent in the AAP and KRC groups than those in the ESH group (Figure 2). No differences in prevalence were observed between the AAP and KRC groups.

**Figure 2.**
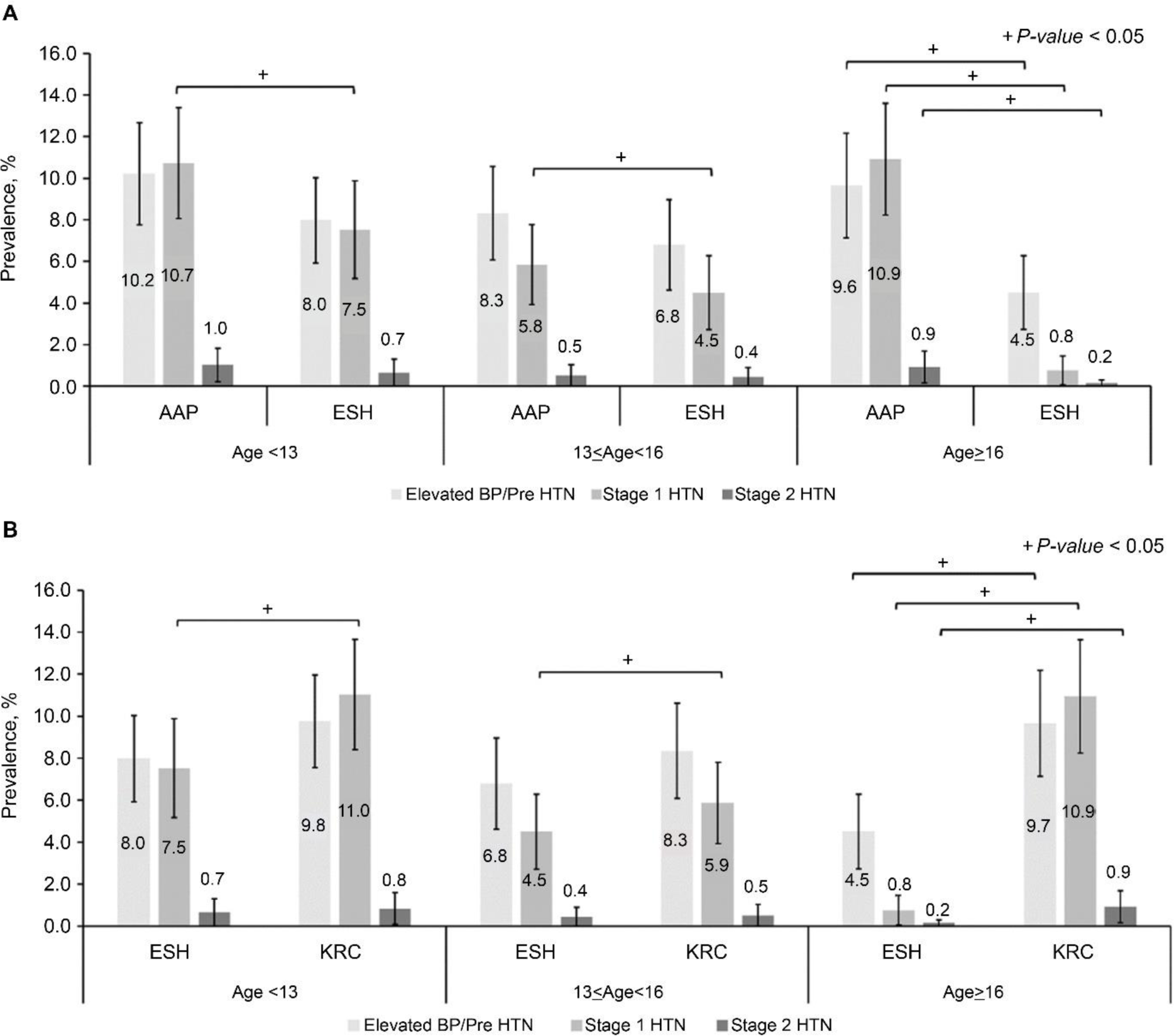
Difference in prevalence of high BP according to age group as classified by A) AAP versus ESH and B) ESH versus KRC. All weighted. AAP, American Academy of Pediatrics; BP, blood pressure; ESH, European Society for Hypertension; HTN, hypertension; KRC, Korea Regional Blood Pressure Classification

### CV Risk Factor Distribution Differences

#### AAP vs. ESH

The AAP reclassified 156 children as having high BP (AAP upwardly reclassified), whereas the ESH reclassified only 11 children (ESH upwardly reclassified). Compared with their matched normotensive counterparts, the AAP upwardly reclassified group showed significant differences in adiposity and metabolic profiles, while the ESH upwardly reclassified group only had lower HDL levels (Table 2).

**Table 2.**
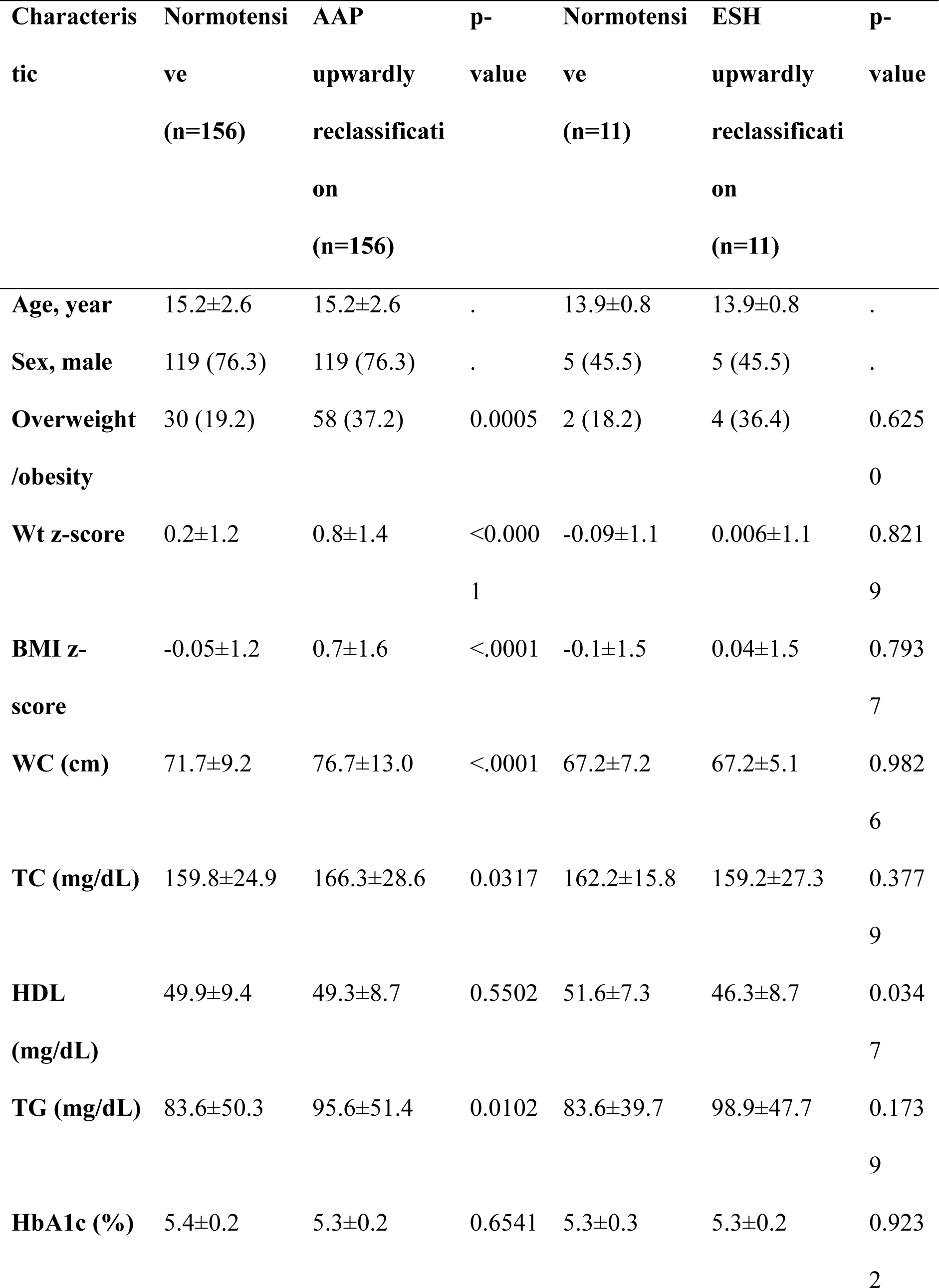
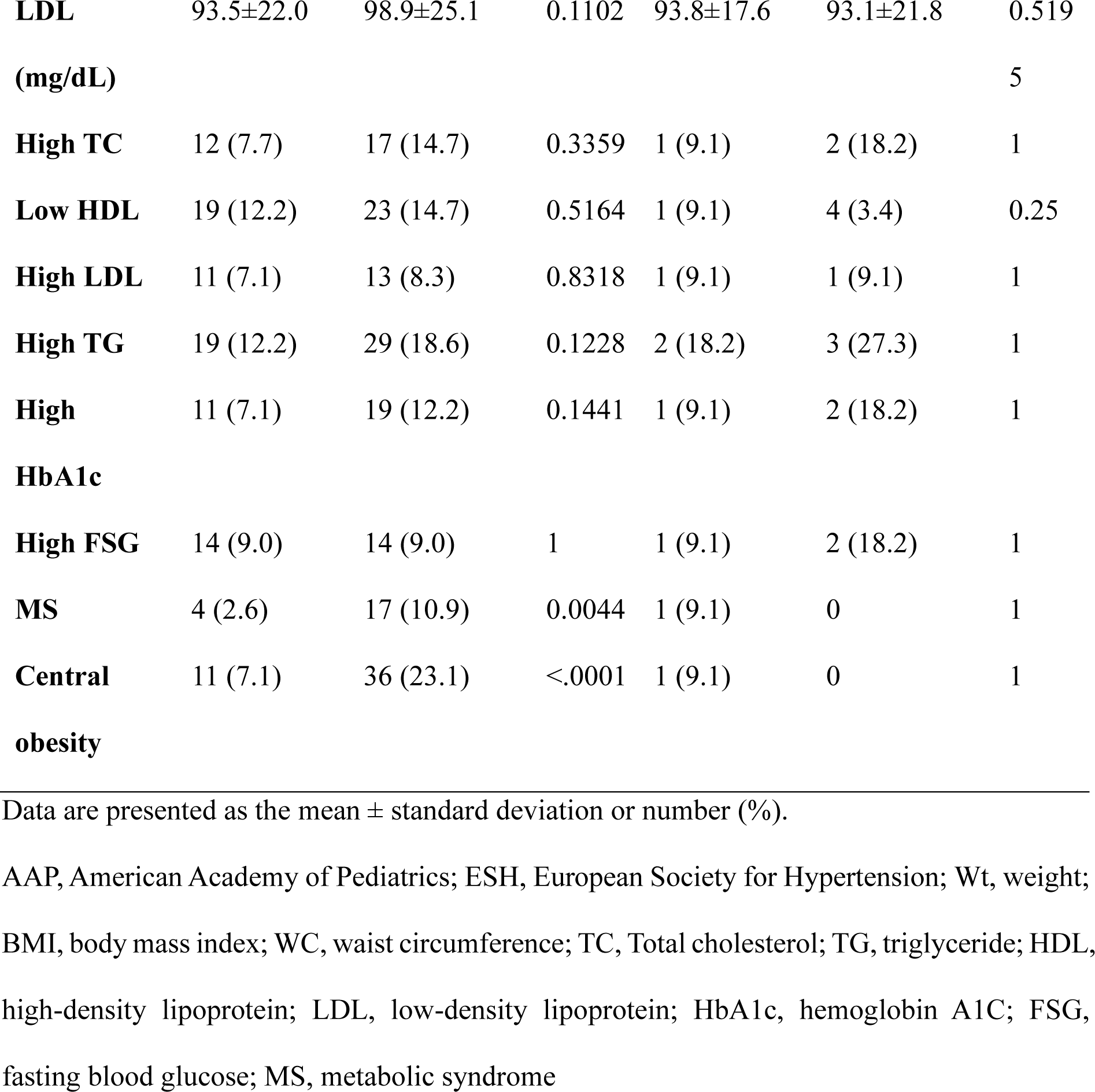
AAP upwardly reclassified and ESH upwardly reclassified vs. age-, sex-, and height-matched consistently normotensive participants, unweighted.

#### AAP vs. KRC

A total of 16 children were reclassified as having high BP by the AAP (AAP upwardly high), while 11 were reclassified by the KRC (KRC upwardly high). No significant differences were observed between the upwardly high group and their normotensive counterparts. However, the KRC classified more non-obese children as having high BP than the AAP (Table 3).

**Table 3.**
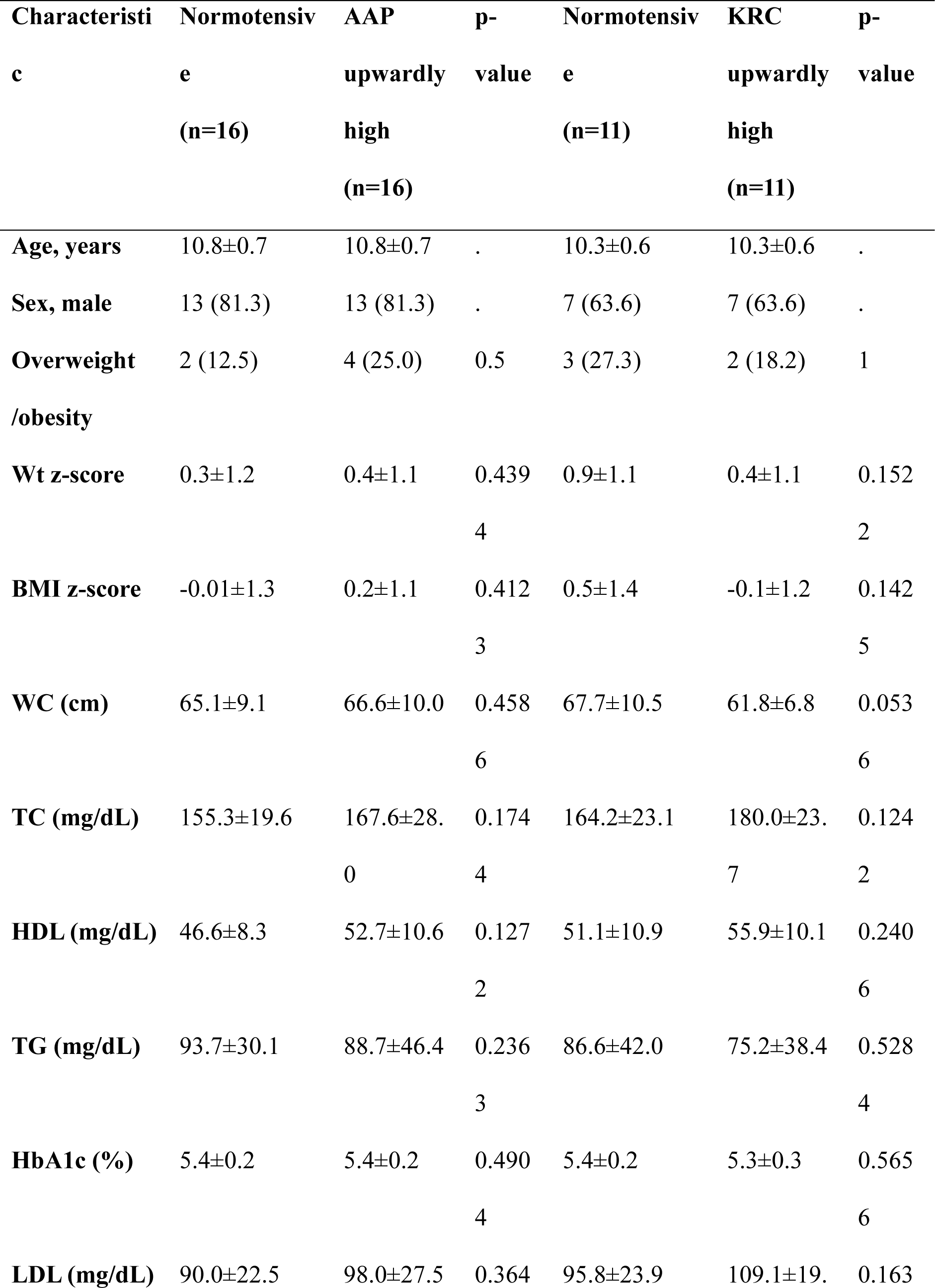
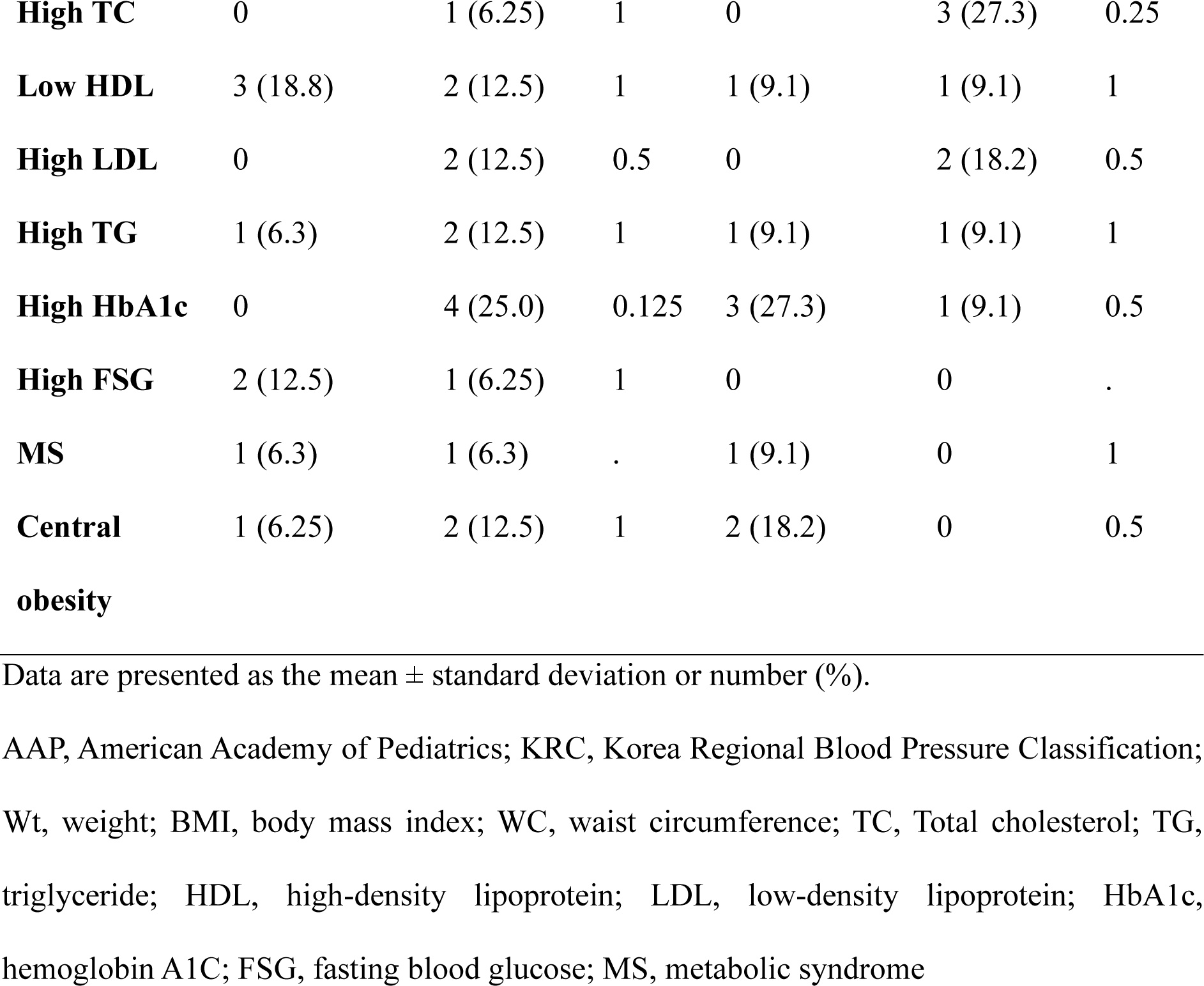
AAP upwardly high and KRC upwardly high vs. age-, sex-, and height-matched consistently normotensive participants, unweighted.

#### ESH vs. KRC

Eleven children were upwardly reclassified as having high BP by the ESH (ESH upwardly high), whereas the KRC reclassified 153 (KRC upwardly reclassified). Compared to their matched normotensive counterparts, ESH upwardly high children showed significant differences in HDL levels, whereas KRC upwardly reclassified children exhibited differences in adiposity and metabolic profiles (Table 4).

**Table 4.**
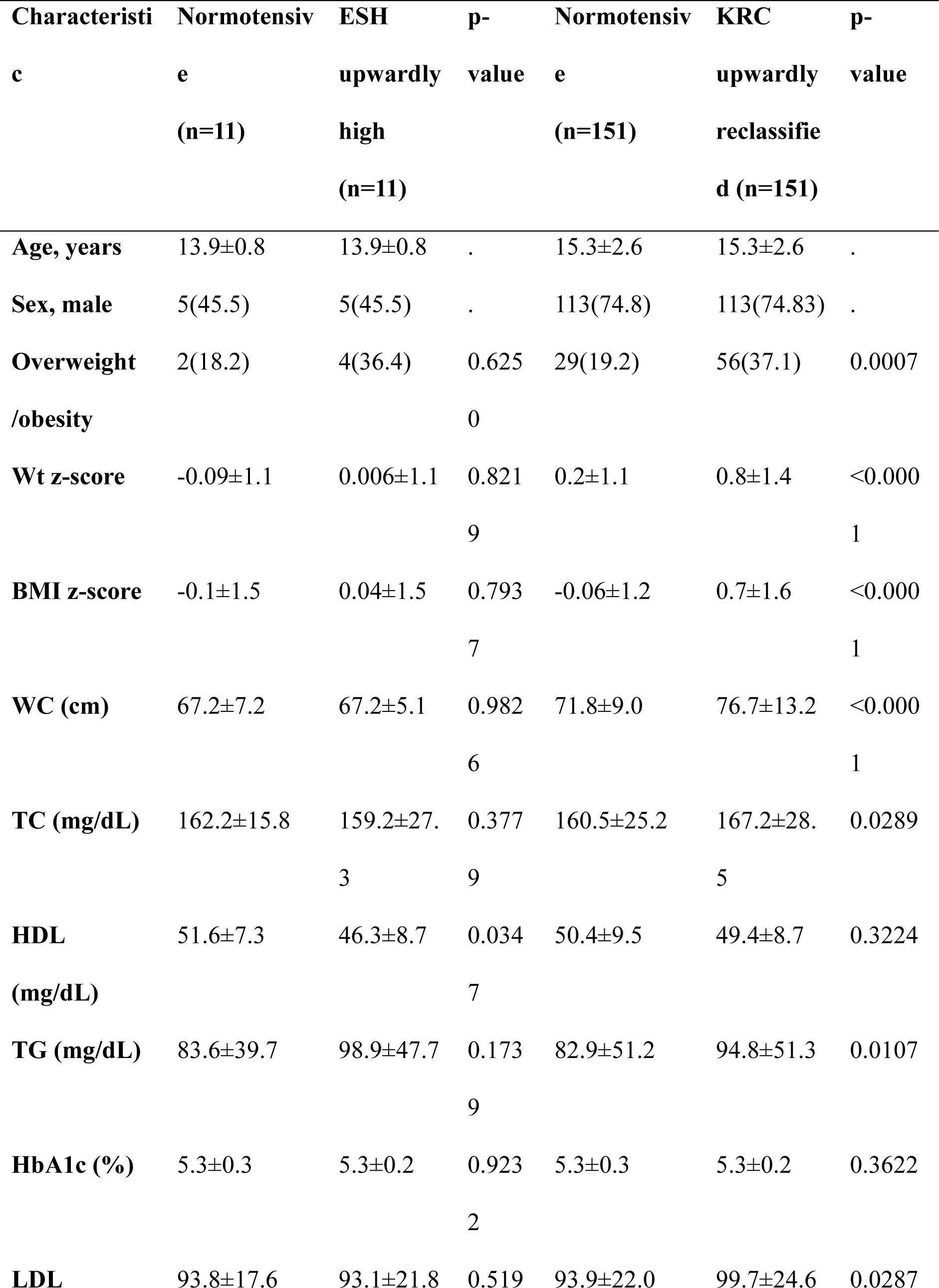
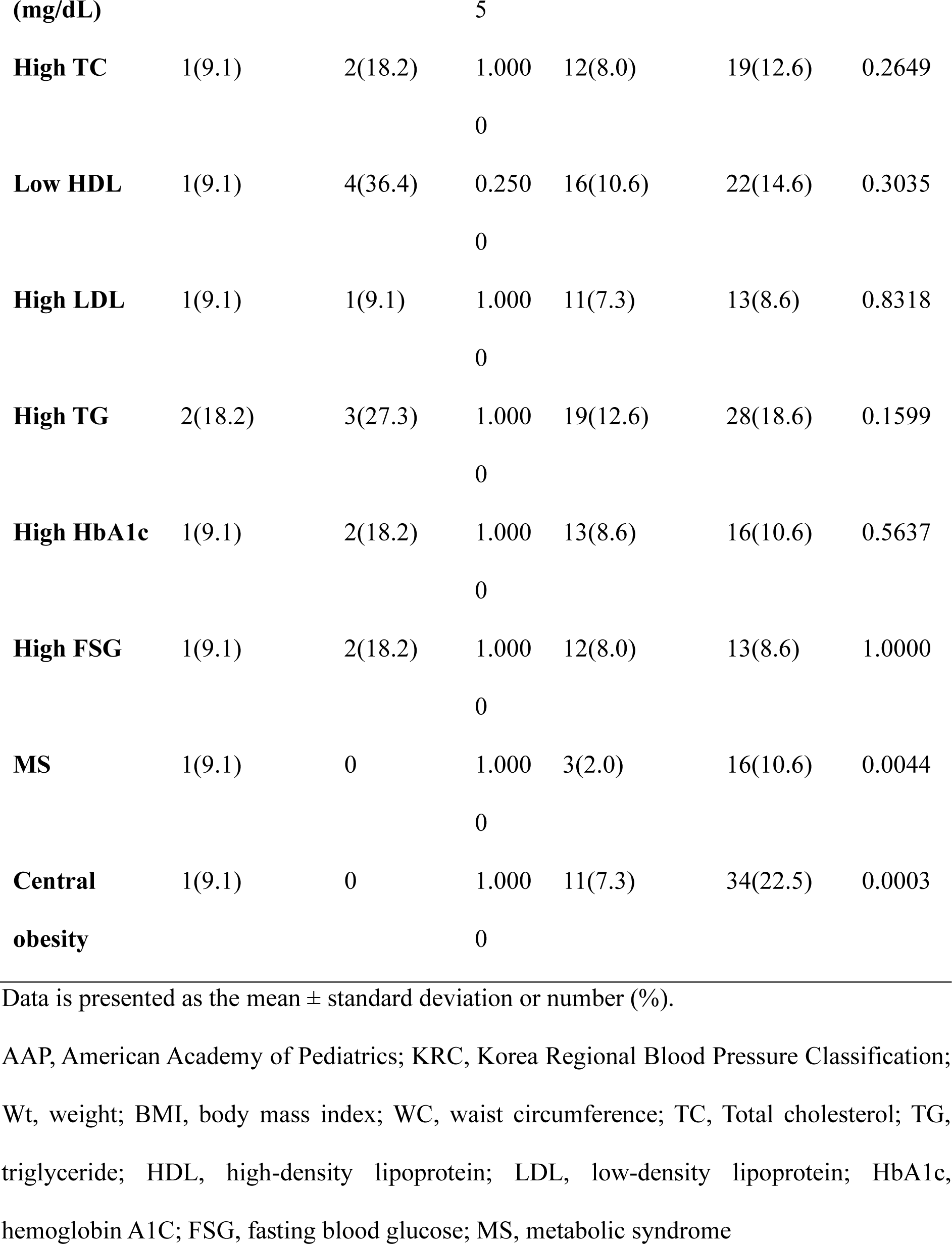
ESH upwardly high and KRC upwardly high vs. age-, sex-, and height-matched consistently normotensive participants, unweighted.

## Discussion

Recently, the prevalence of HTN in Korean children has increased.^31^ We sought to compare the clinical characteristics across different BP categories, aiming to propose uniform and practically applicable classification criteria tailored for Koreans. Notable differences include the AAP’s utilization of a lower BP reference value, excluding obese children, and applying a static cutoff BP value at a younger age (13 years) than the ESH (16 years), with lower static cutoff values.^23,24^

Previous studies have consistently shown a higher prevalence of high BP in the AAP compared to that in the ESH,^32–37^ consistent with the findings in our study. There was a concern regarding a potential underdiagnosis of HTN in the AAP for youths older than 13 years, as a static cutoff BP value derived from adulthood HTN definitions was applied. The findings support the applicability of the AAP in Korean teenagers (13-15 years); only stage 1 HTN increased, with no significant decrease. Chinese^38^ and Spanish^33^ studies showed similar trends, suggesting that the AAP could simplify diagnostic criteria and enhance real-world HTN detection. Because BP can be influenced by race, ethnicity, and obesity, several countries have established local BP references.^39–42^ Conversely, international BP reference values are derived from nationally representative cross-sectional surveys.^43^ Studies comparing local and international references as well as the inclusion/exclusion of overweight or obese children’s BP data have revealed significant discrepancies in high BP prevalence across different reference tables.^44–49^ A study conducted by Fan et al.^48^ in China applied a local reference table, the AAP, the fourth report (a previous version of the BP guidelines before the AAP 2017), and international BP reference tables to a longitudinally followed cohort. This study found that high BP, as defined by the AAP and local BP references, effectively predicted subclinical CVD in adulthood. From this perspective, we applied a normal-weight Korean BP reference to the AAP BP classification (KRC).

We focused on the characteristics of children reclassified from normotensive to high BP groups using various criteria. Prior research has explored the clinical differences in defining high BP using the AAP and ESH, leading to controversies regarding their ability to detect CVD risks. A study by Di Bonito et al.^34^ showed that despite no prevalence difference in LVH and relative wall thickness between individuals with high BP defined by the AAP and ESH, the AAP identified more abnormal left ventricular geometries among Italian overweight/obese children. Another study by Di Bonito et al.^37^ found that Italian overweight/obese children reclassified as having high BP by the AAP but normotensive by the ESH had a higher BMI, poorer metabolic profiles, and higher left ventricular mass index than consistently normotensive children.

Additionally, Kim et al.^50^ reported a difference in the prevalence of BP according to the AAP and ESH in Korean youth. They showed a higher prevalence of high BP in patients with the AAP than in patients with the ESH. Those who were newly diagnosed with high BP by the AAP were more obese and had more severe cardiometabolic risk factors than normotensive subjects. In contrast, a study by Antolini et al.^32^ reported that although the AAP increased the prevalence of high BP relative to the ESH, they did not enhance the identification of early cardiac organ damage when weight was adjusted in the analyses. Some studies compared the AAP with the fourth report. Sharma et al.^51^ and Yang et al.^52^ analyzed the clinical characteristics of children newly diagnosed with high BP or upwardly reclassified with a higher stage BP by the AAP, comparing them with age-, sex-, and height-matched normotensive children according to both guidelines. The upwardly reclassified children displayed adverse lipid profiles and a cluster of CV risk factors compared with matched normotensive children. These findings align with our study’s comparison of the AAP and ESH (the ESH shares the BP reference table from the fourth report).^24^

Children newly classified as having high BP under the KRC were primarily reclassified based on their DBP. Additionally, there was a tendency for non-obese children to be reclassified into the high-BP group with the KRC. Increased DBP in non-obese children has been associated with insulin resistance in young adults and metabolic disorders.^53^ This suggests the importance of using local reference BP tables. A comparison of the KRC with the ESH revealed similar patterns in the prevalence of CVD risks to the AAP and ESH comparisons. This emphasizes that high BP determined by the AAP and KRC identifies more children with CVD risk factors than high BP determined by the ESH, potentially leading to overdiagnosis of high BP and heightened sensitivity for CVD risk factors.

Both the AAP and ESH guidelines recommend lifestyle modifications for the initial management of high BP in children.^23,24^ We suggest that the KRC is more effective in identifying individuals at risk of CVD and this will contribute to the overall awareness and management of CV health in the pediatric population in Korea.

Our study had several strengths. First, we used national survey data from the KNHANES, which provides a fully representative sample of the Korean pediatric population. The use of comprehensive data strengthens the validity and applicability of the study to a broader population. Second, this study is the first to compare different BP guidelines and the impact of applying normal-weight Korean BP references in Korean youth, with the aim of determining the most reliable BP classification criteria for risk control. However, this study had certain limitations. First, the number of children reclassified according to each guideline was small, potentially affecting the statistical power and generalizability of the findings. Second, the reliance on cross-sectional data from the KNHANES, which includes only one BP measurement, could affect the real-world prevalence of high BP. Third, the retrospective cross-sectional nature of this study prevents assessment of the impact of high BP, including potential TOD.

In conclusion, the KRC effectively identified Korean youth with a higher prevalence of CVD risk factors, categorizing them as having a high BP. Contrary to concerns about underdiagnosis, applying a static HTN definition starting at the age of 13 years in Korean youths did not result in an underdiagnosis of high BP.

### Perspectives

Using the KRC as a uniform and simplified set of BP criteria can reduce the likelihood of diagnosis failure and enhance early CVD risk control. Further research is warranted to investigate the longitudinal impact of KRC use on the actual CVD outcomes in adulthood in the Korean pediatric population.

## Data Availability

The datasets generated and/or analysed during the current study are available in the KNHANES webpage repository, (https://knhanes.kdca.go.kr/knhanes/main.do) with the permission of the KDCA.

## Non-standard Abbreviations and Acronyms

AAP: American Academy of Pediatrics
ESH: European Society for Hypertension
KDCA: Korea Disease Control and Prevention Agency
KNHANES: Korean National Health and Nutritional Examination Survey
KRC: Korea Regional Blood Pressure Classification
MS: metabolic syndrome
TOD: target organ damage

## Novelty and Relevance

### What Is New?

- We developed the Korea Regional Blood Pressure (BP) Classification (KRC).

### What Is Relevant?

- The KRC effectively identified Korean youth with a higher prevalence of cardiovascular disease (CVD) risk factors.
- Applying a static hypertension definition starting at the age of 13 years in Korean youths did not result in an underdiagnosis of high BP.

### Clinical/Pathophysiological Implications?

- Using the KRC as a uniform and simplified set of BP criteria can reduce the likelihood of diagnosis failure and enhance early CVD risk control.

## Acknowledgements

None

## Supplemental Material

Tables S1-S4 Figure S1

---

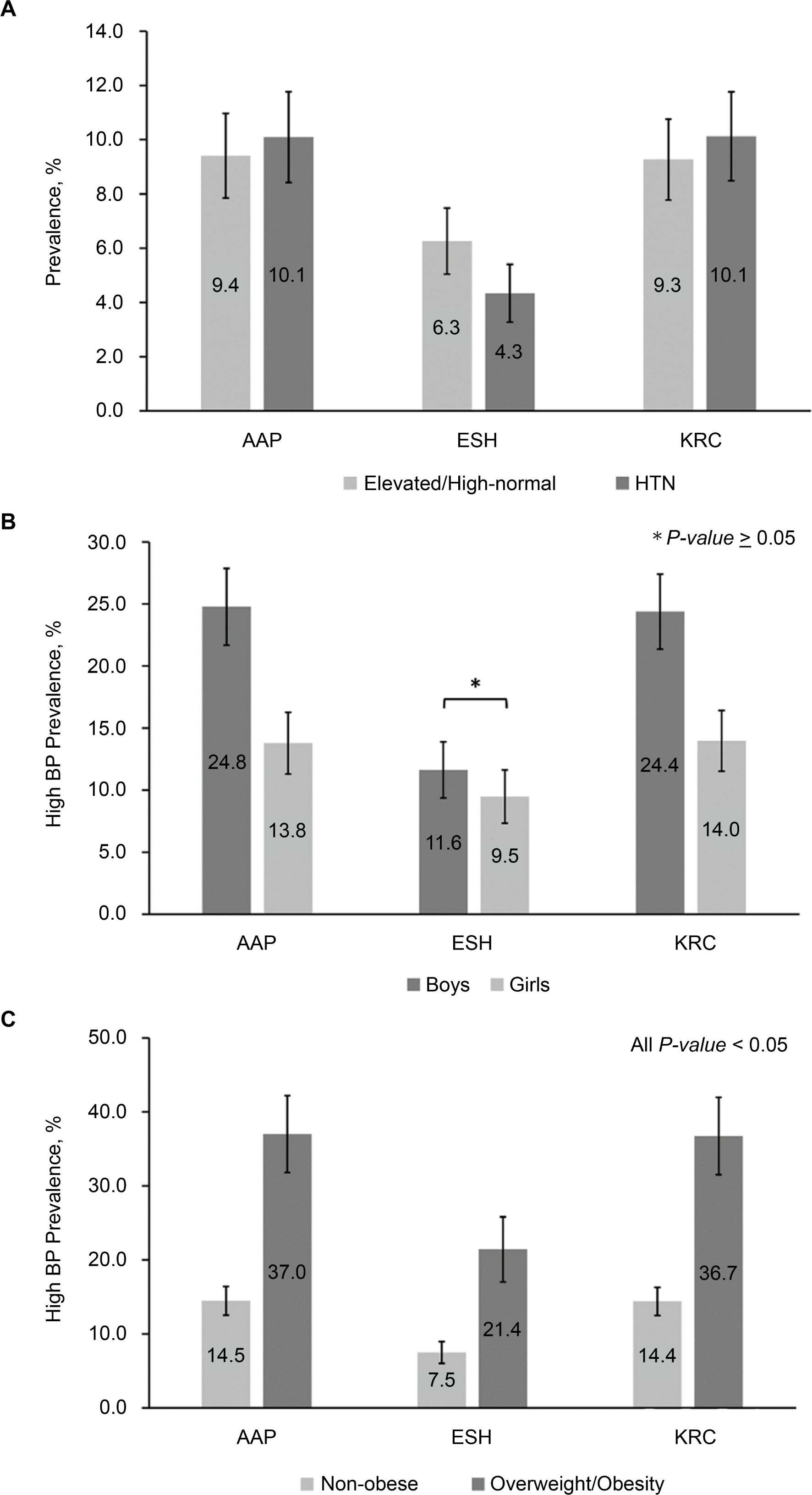

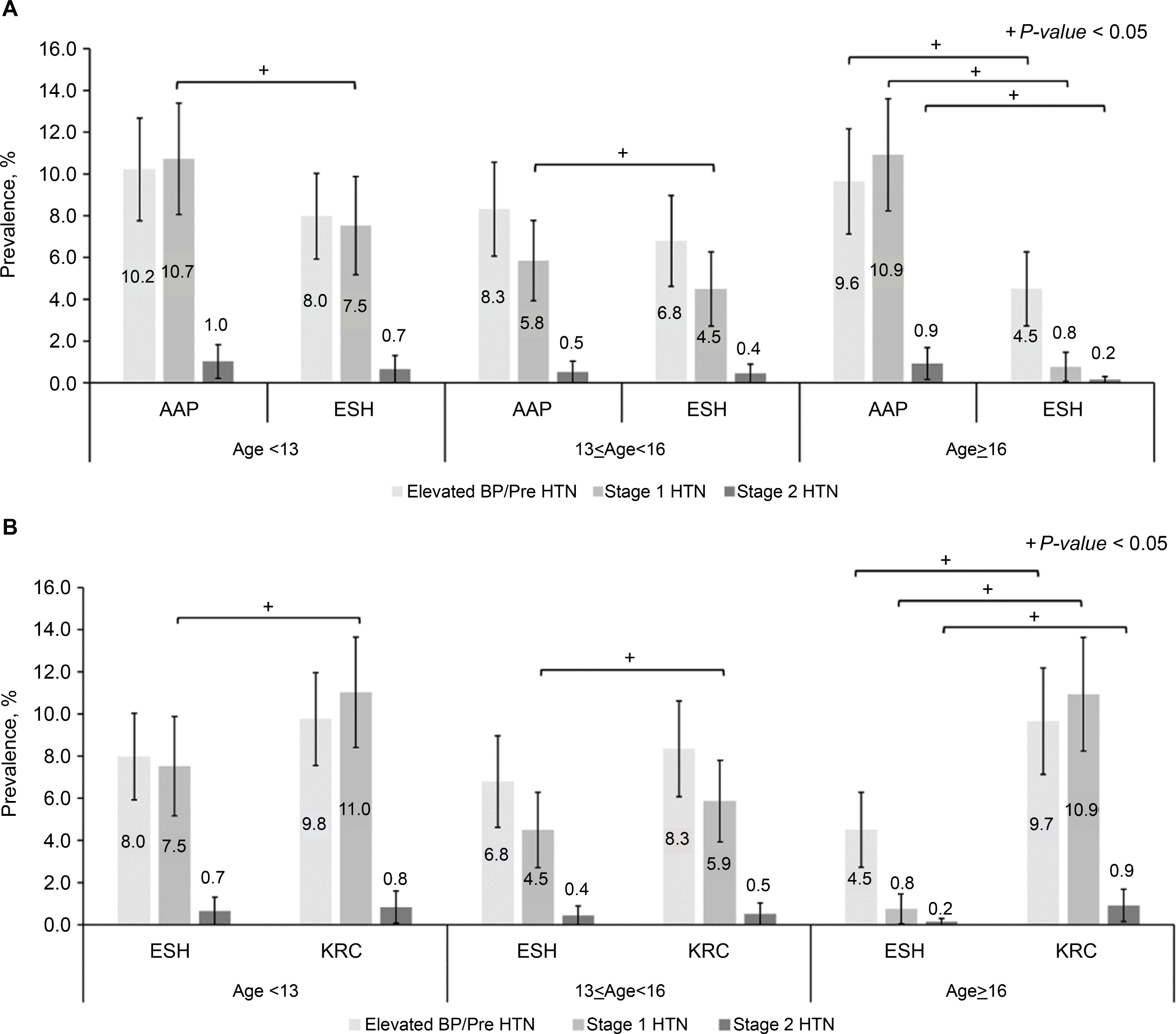

